# Divergence of wastewater SARS-CoV-2 and reported laboratory-confirmed COVID-19 incident case data coincident with wide-spread availability of at-home COVID-19 antigen tests

**DOI:** 10.1101/2023.02.09.23285716

**Authors:** Alexandria B. Boehm, Marlene K. Wolfe, Bradley J. White, Bridgette Hughes, Dorothea Duong

## Abstract

Concentrations of SARS-CoV-2 RNA in wastewater settled solids from publicly owned treatment works (POTWs) historically correlated strongly with laboratory confirmed incident COVID-19 case data. With the increased availability of at-home antigen tests since late 2021 and early 2022, laboratory test availability and test seeking behavior has decreased. In the United States, the results from at-home antigen tests are not typically reportable to public health agencies and thus are not counted in case reports. As a result, the number of reported laboratory-confirmed incident COVID-19 cases has decreased dramatically, even during times of increased test positivity rates and wastewater concentrations of SARS-CoV-2 RNA. Herein, we tested whether the correlative relationship between wastewater concentrations of SARS-CoV-2 RNA and reported laboratory-confirmed COVID-19 incidence rate has changed since 1 May 2022, a point in time immediately before the onset of the BA.2/BA.5 surge, the first surge to begin after at-home antigen test availability was high in the region. We used daily data from three POTWs in the Greater San Francisco Bay Area of California, USA for the analysis. We found that although there is a significant positive association between wastewater measurements and incident rate data collected after 1 May 2022, the parameters describing the relationship are different than those describing the relationship between the data collected prior to 1 May 2022. If laboratory test seeking or availability continues to change, the relationship between wastewater and reported case data will continue to change. Results suggests that, assuming SARS-CoV-2 RNA shedding remains relatively stable among those infected with the virus as different variants emerge, that wastewater concentrations of SARS-CoV-2 RNA can be used to estimate COVID-19 cases as they would have been during the time when laboratory testing availability and test seeking behavior were at a high (here, before 1 May 2022) using the historical relationship between SARS-CoV-2 RNA and COVID-19 case data.

## Introduction

Concentrations of genetic material from various viruses in wastewater solids collected from publicly owned treatment works (POTWs) correlate well with clinical testing data on disease occurrence^1–3^. SARS-CoV-2 RNA concentrations in wastewater solids have been shown to correlate strongly with laboratory-confirmed cases of COVID-19^4–6^. Similarly, concentrations of influenza A and B, RSV, metapneumovirus, parainfluenza, rhinovirus, seasonal coronaviruses, and mpox nucleic acids in POTW wastewater solids correlate well with clinical sample percent positivity rates and/or reported disease incidence in the contributing communities^2,3,7–9^.

Wastewater is a less biased approach than clinical testing for gaining insight to community health as it represents a composite biological sample containing contributions from the entire contributing population. Human excretions including feces, urine, sputum, saliva, and mucus are present in wastewater. Viruses and their genetic material adsorb to solids in the wastestream and as a result, their concentrations are enriched orders of magnitude in the solids compared to the liquid phase of wastewater ^5,9–12^. Therefore, solids represent a natural concentrator of virus in wastewater and an ideal matrix for measuring their concentrations for wastewater-based epidemiology.

Our and others’ previous work showed strong correlations between SARS-CoV-2 RNA concentrations in wastewater and laboratory-confirmed COVID-19 case data ^13–18^. This work was primarily completed before the emergence of the Omicron variant, and used COVID-19 case data compiled by public health departments, almost entirely reliant on testing of patient specimens performed in clinical laboratories. The emergence of Omicron occurred at the same time at-home antigen tests were first made available by the United States federal government free of charge on 16 January 2022 (at the height of the Omicron BA.1 surge)^19^. The results of at-home antigen tests are not typically reportable to public health agencies so their positive results are not recorded in public health databases^20^.

The goal of the present study is to compare the relationship between SARS-CoV-2 RNA concentrations measured in POTW wastewater solids to reported laboratory-confirmed COVID-19 incident cases before and after at-home antigen tests were widely available. Daily concentrations of SARS-CoV-2 from three large POTWs located in the Greater San Francisco Bay Area of California, USA are used for the analysis, along with publicly available data on incident laboratory-confirmed COVID-19 cases and laboratory test positivity rates.

## Methods

The study was reviewed by the IRB at Stanford University and the IRB has determined that this research does not involve human subjects as defined in 45 CFR 46.102(f) and therefore does not require IRB oversight.

### Study sites

Daily samples between 1 December 2020 and 16 January 2023 of wastewater settled solids were obtained from three publicly owned treatment works (POTWs) San Jose (SJ) Oceanside (OS), and Sacramento (SAC) serving populations in Santa Clara County, San Francisco County, and Sacramento County, California USA, respectively. The plants serve 1.5 million, 250,000, and 1.5 million people, respectively, representing 77%, 30%, and 93% of their county populations, respectively. Detailed methods for sample collection have been presented elsewhere^14,21–24^. In brief, grab samples of solids were collected from the primary clarifier daily in sterile containers and placed at 4°C and couriered to the laboratory where they were processed immediately. A total of 777, 737, and 769 samples were included in this study from SJ, OS, and SAC, respectively. Note that a subset of these data have already been published; in particular data from all three POTWs collected between 1 December 2020 and 31 March 2021 was previously published by Wolfe et al.^14^ and data between 1 January 2022 and 13 April 2022 were published by Boehm et al.^25^. Otherwise, the data presented herein has not been previously published.

### Laboratory analyses

Samples were processed the same day they were collected in the laboratory to measure concentrations of SARS-CoV-2 N gene and an internal endogenous control pepper mild mottle virus (PMMoV). The methods are detailed in other peer-reviewed publications^14,21^ and available on protocols.io^21–23^. The N gene target is conserved across SARS-CoV-2 variants to date. In brief, the samples were centrifuged to dewater the solids, resuspended in a buffer spiked with an exogenous viral control (bovine coronavirus, BCoV), homogenized, and then centrifuged. Nucleic-acids were extracted and purified from the supernatant using a commercial kit and then the RNA extract was subjected to an inhibitor removal step. The resultant RNA was used immediately as template in reverse-transcription polymerase chain reactions (RT-PCR) in a digital droplet format to measure the SARS-CoV-2 N gene, PMMoV, and BCoV. Positive and negative extraction and RT-PCR controls were included in all runs, and we ensured that BCoV recovery was greater than 10% in all samples. Previous work showed that inhibition using this method was not present^24^. Results for SARS-CoV-2 N gene and PMMoV are reported as copies of target per dry weight of solids. Wastewater data are available through the Stanford Digital Repository (https://doi.org/10.25740/xy132dg9314).

### Case data

Publicly available data on the laboratory-confirmed incidence rate and test positivity rate for Santa Clara, San Francisco, and Sacramento Counties are available online (https://data.chhs.ca.gov/dataset/covid-19-time-series-metrics-by-county-and-state). Incidence rates for populations served by specific POTWS (for each sewershed) were provided by California Department of Public Health. Incidence and positivity rate data are reported as a function of the “episode date” which is the earliest reported date associated with the specimen (symptom onset, collection, hospitalization, or death; but usually specimen collection date) and include results of nucleic-acid amplification tests (hereafter, “PCR tests”). Seven-day smoothed incidence and positivity rate data are used to represent incidence and positivity rate hereafter, because it is well understood that these data have substantial day-of-week effects^26^.

### Statistical analysis

We used Pearson’s r to test for linear relationships between sewershed-aggregated and county-aggregated incidence rate data for each POTW (three tests total). We used Kendall’s tau to test for association between county-aggregated positivity and incidence rates for each POTW (three tests). We used Kendall’s tau to test for association between wastewater and clinical case data sets. We tested for associations between measured N, N/PMMoV, 5-d trimmed average N, and 5-d trimmed N/PMMoV, and incidence rate and positivity rates for each POTW resulting in twenty-four Kendall’s tau tests. We chose Kendall’s tau as it does not require that data are normally distributed and therefore did not require data transformation; however, results are unchanged if other tests of association are used.

We tested whether the relationship between wastewater measurements of SARS-CoV-2 RNA and incidence rate differed for data collected before, and after 1 May 2022 using the following linear model:

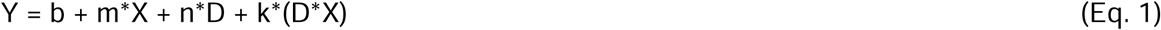

where Y is log_10_-transformed 7-day smoothed incidence rate data aggregated at the county where the POTW is located, X is the log_10_-transformed 5-d trimmed average N/PMMoV, D is a categorical, binary variable indicating whether the date is after 1 May 2022 (D = 0 if the date is before 1 May 2022, and 1 if it is on or after this date), b is the intercept, m is the coefficient on X, n is the coefficient on D, and k is the coefficient on the interaction term D*X. We chose 1 May 2022 as the date to bifurcate the data since this date occurs at the onset of the BA.2/BA.5 COVID-19 case surge in this region. This represents the first surge that began after at-home antigen tests became widely available in the region. We confirmed via visual examination that this date represented approximately when the relationship between incidence rate and wastewater concentrations appeared to change (see results).

The model was applied to each sewershed (model was run three times). We chose to use the 5-d trimmed average wastewater concentration as it eliminates the influence of intrinsic variability associated with measurements from a complex, environmental matrix; and we chose to use N/PMMoV as PMMoV normalization can control for variation in RNA extraction efficiency and fecal strength of the solids^14,27,28^. However, we explored whether using raw data (not 5-d trimmed averages) and unnormalized N gene concentrations as X in the model, and results were generally unchanged.

Using Eq. 1, we tested whether the relationship between positivity and incidence rate was different for data collected before, and after 1 May 2022. In that application, Y = log_10_-transformed incidence rate, and X = log_10_-transformed positivity rate. The model was applied to each sewershed (model was run 3 times).

We confirmed the Eq. 1 model residuals were approximately normal. All statistical analyses were carried out in R studio (Version 1.4.1106) using R (Version 4.0.5). We used alpha = 0.05 which corresponds to a p value of 0.0013 with a Bonferroni correction (0.05/36 tests).

## Results

Positive and negative RT-PCR and nucleic-acid extraction controls were positive and negative, respectively, indicating assays performed well and there was no contamination during sample pre-analytical and analytical processing. BCoV recovery was greater than 10% (median across samples from each POTW was 80%), and PMMoV concentrations were fairly consistent across samples (median ± interquartile range of 9.0×10^8^±4.3×10^8^ cp/g (SAC), 5.2×10^8^±3.4×10^8^ cp/g (OS), and 1.5×10^9^±6.0×10^8^ cp/g (SJ)) indicating reasonable RNA extraction efficiency and fairly consistent fecal strength of wastewater (Figure S1). We previously published details related to the dMIQE^29^ and EMMI^30^ guidelines for these measurements^14,25^.

Sewershed-aggregated incidence rates correlate strongly with county-level incidence rates (Pearson’s r of 1.00 for SJ, 0.99 for OS, and 0.93 for SAC, p<10^−15^ for all). We use county-aggregated incidence rate hereafter to represent sewershed-aggregated incidence rate given their high correlations, because the county-aggregated data are publicly available and can be shared readily. Positivity rate aggregated at the sewershed is not available, so we used county-aggregated positivity rate as a proxy for the sewershed given the high degree of correlation between county and sewershed-aggregated incidence rate.

Across all three sewersheds, SARS-CoV-2 N gene concentrations, and those concentrations normalized by PMMoV (raw and 5-day trimmed averages) were positively and significantly correlated to county-aggregated COVID-19 incidence rate, and county-aggregated positivity rate (Kendall’s tau between 0.51 and 0.62, p<10^−15^ for correlations with incidence rate, and Kendall’s tau between 0.68 and 0.80, p<10^−15^ for correlations with positivity rate), consistent with previously published work^4,6,17,31^. The ranges reported in the previous sentence include tau calculated with the four considered wastewater variables for the three plants.

County-aggregated incidence rate and positivity rate are well correlated (Kendall’s tau 0.79 for SAC, 0.96 for SJ, 0.88 for OS, all p<10^−15^), but there is a change in their relationship starting approximately 1 May 2022, at the onset of the BA.2/BA.5 surge (Figure S2). A multiple regression model was used to test whether the relationship between positivity rate and incidence rate was significantly different before and after 1 May 2022 at each of the considered counties, and those models indicated that the relationship was significantly different before and after 1 May 2022 with different slope and/or intercepts (Table S1).

When log_10_-transformed incidence rate and positivity rate are plotted against log_10_-transformed, 5-d trimmed average wastewater concentrations of SARS-CoV-2 N gene normalized by PMMoV at the three sewersheds, the data generally fall on a single line (Figures 1 - 3). However, data collected starting approximately 1 May 2022 fall off the line represented by data collected before 1 May 2022 on the incidence rate - wastewater plot; the same pattern is not evident on the positivity rate - wastewater plot (Figures 1-3). The relationship between log_10_-transformed incidence rate and log_10_-transformed N/PMMoV concentrations collected after 1 May 2022 appears approximately linear, but it is visually offset from earlier measurements.

**Figure 1.**
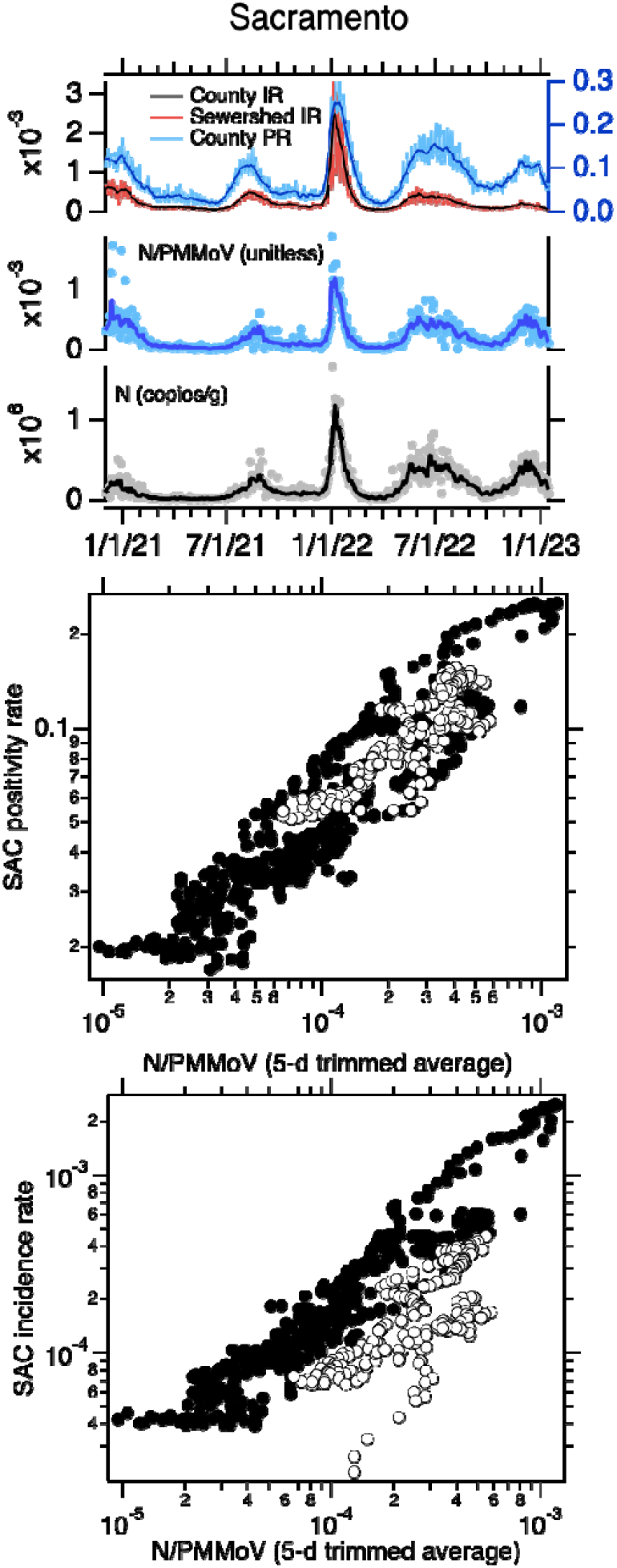
Top panel. Sacramento (SAC) time series of concentrations of N and N/PMMoV in wastewater, incidence rates, and positivity rates between 1 December 2020 until 16 January 2023. The solid lines represent 5-d trimmed averages for the wastewater data, and 7-d moving averages for the incidence and positivity rates. The county-aggregated and sewershed-aggregated incidence rate data fall almost directly on top of each other, obscuring the view of both. The scales for incidence and positivity rates are on the left and right axes, respectively. Middle and bottom panels. County-aggregated (Sacramento County, SAC) positivity rate and incidence rate versus 5-d trimmed N/PMMoV; white symbols are data collected on and after 1 May 2022.

**Figure 2.**
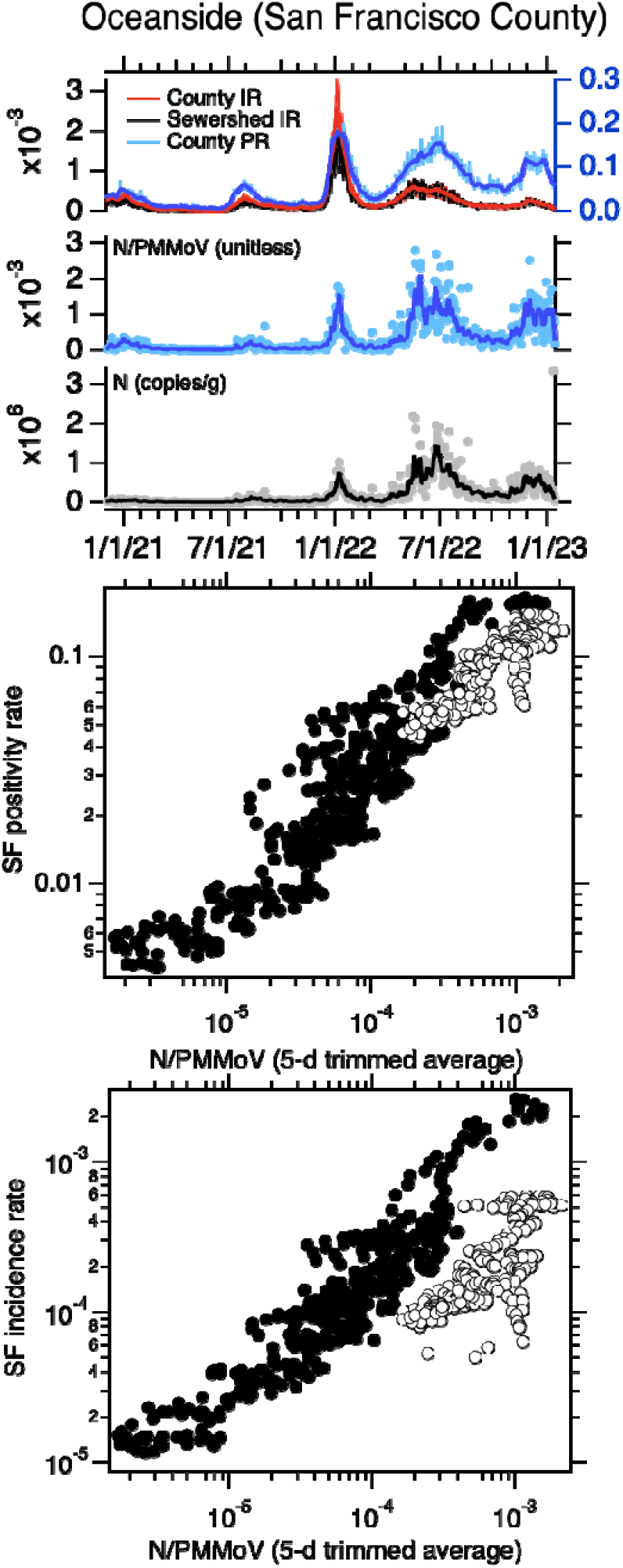
Top panel. Oceanside (OS) time series of concentrations of N and N/PMMoV in wastewater, incidence rates, and positivity rates between 1 December 2020 until 16 January 2023. The solid lines represent 5-d trimmed averages for the wastewater data, and 7-d moving averages for the incidence and positivity rates. The county-aggregated and sewershed-aggregated incidence rate data fall almost directly on top of each other, obscuring the view of both. The scales for incidence and positivity rates are on the left and right axes, respectively. Middle and bottom panels. County-aggregated (San Francisco County, SF) positivity rate and incidence rate versus 5-d trimmed N/PMMoV; white symbols are data collected on and after 1 May 2022.

**Figure 3.**
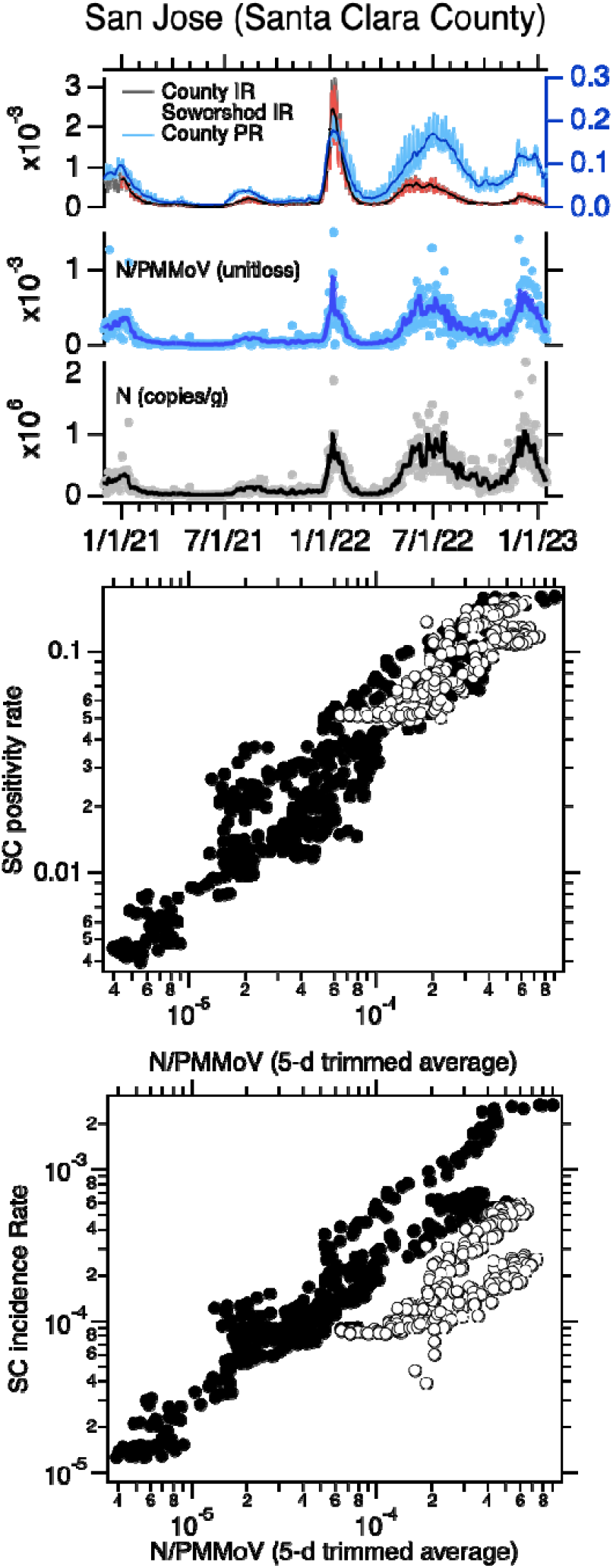
Top panel. San Jose (SJ) time series of concentrations of N and N/PMMoV in wastewater, incidence rates, and positivity rates between 1 December 2020 until 16 January 2023. The solid lines represent 5-d trimmed averages for the wastewater data, and 7-d moving averages for the incidence and positivity rates. The county-aggregated and sewershed-aggregated incidence rate data fall almost directly on top of each other, obscuring the view of both. The scales for incidence and positivity rates are on the left and right axes, respectively. Middle and bottom panels. County-aggregated (Santa Clara County, SC) Positivity rate and incidence rate versus 5-d trimmed N/PMMoV; white symbols are data collected on and after 1 May 2022.

Using multiple linear regression, we tested whether the linear relationship between log_10_-transformed incidence rate and log_10_-transformed 5-d trimmed average N/PMMoV is the same for data collected before and after 1 May 2022. All models were statistically significant with adjusted R^2^ values between 0.81 and 0.86 (p<10^−15^). For SAC, the slope of the relationship between the two variables is not different for data collected before and after 1 May 2022 (slope = 0.88) but the intercept is different (−0.26 compared to -0.87 for data collected before and after 1 May 2022, respectively). For SJ, the slope of the relationship between the two variables is not different for data collected before and after 1 May 2022 (slope = 0.96); but the intercept is different (0.29 compared to -0.66 before and after 1 May 2022, respectively). For OS, both the slope and the intercept of the relationship between incidence rate and wastewater are different for data collected before and after 1 May 2022 (slope = 0.65 and 0.35 for data before and after 1 May 2022, respectively; intercept = -1.2 compared to -2.73 before and after 1 May 2022, respectively). Results reported here are summarized in Table 1. We repeated the modeling using N (both raw and 5-d trimmed averaged data) and raw N/PMMoV as the dependent variables (X in equation 1), and results were similar.

**Table 1.** Coefficients for the model Y = b + m*X + n*D + k*(D*X) presented as equation 1 in the text where X is log_10_-transformed 5-d trimmed N/PMMoV and Y is log_10_-transformed incidence rate. Adjusted R-square values and p-values for the model are provided, as well as the F statistic (and the degrees of freedom, DF). Coefficient values with standard errors are reported as well as p-values in parentheses. With Bonferroni corrections, p must be less than 0.0013 for alpha = 0.05. If n is significantly different from 0, then the intercept for the linear relationship between X and Y before and after 1 May 2022 is different and equal to b + n. If k is significantly different from 0, then the slope for the linear relationship between X and Y is different before and after 1 May 2022 and equal to m + k.

| POTW | b | m | n | k | Adjusted $R^2$ | F-statistic |
| --- | --- | --- | --- | --- | --- | --- |
| SJ | 0.29±0.07<br>($1.3 \times 10^{-5}$) | 0.96±0.02<br>($<10^{-100}$) | -0.96±0.18<br>($<10^{-7}$) | -0.12±0.05<br>(0.014) | 0.86 ($10^{-15}$) | 1545 on 3<br>and 773 DF |
| OS | -1.22±0.05<br>($<10^{-100}$) | 0.65±0.01<br>($<10^{-100}$) | -1.51±0.22<br>($<10^{-11}$) | -0.29 ±0.06<br>($<10^{-5}$) | 0.81<br>($<10^{-15}$) | 1070 on 3<br>and 733 DF |
| SAC | -0.26±0.06<br>($2.9 \times 10^{-5}$) | 0.88±0.02<br>($<10^{-100}$) | -0.61±0.15<br>($6.7 \times 10^{-5}$) | -0.067±0.04<br>(0.10) | 0.83<br>($<10^{-15}$) | 1270 on 3<br>and 765 DF |

As equation 1 describes the relationship between log_10_-transformed incident rates and 5-d trimmed N/PMMoV well, it implies that incident rate and N/PMMoV are related by a power law relationship where incident rate = 10^intercept^(N/PMMoV)^slope^.

## Discussion

In the Greater San Francisco Bay Area of California, USA, the BA.2/BA.5 COVID-19 case surge began around 1 May 2022 and represents the first surge in the region to begin after at-home antigen tests became widely available. The relationship between wastewater concentrations of SARS-CoV-2 RNA and laboratory-confirmed incidence rates of COVID-19 in the contributing community changed after 1 May 2022, likely a result of the change in regional PCR test seeking behaviors and testing availability. Because case data is likely underestimating the true cases in the community, this divergence indicates that wastewater may be particularly useful for obtaining a less biased estimate of community infection.

The relationship between SARS-CoV-2 RNA concentrations and laboratory-confirmed incident cases has previously been described as a power-law relationship (i.e., it is linear on a log-log plot)^1^. We found that data collected both before and after 1 May 2022 at the three POTWs follow power-law relationships albeit with different parameter values. In particular, we found the slope of the log-log relationship to be similar for data collected before and after 1 May 2022 for two of the three POTWs, but the intercept to be different at all three. At all three POTWs, the intercept is smaller for data collected after 1 May 2022. This can be interpreted to mean that for data considered after 1 May 2022, that a one log-change in wastewater concentration indicates a log-change (between 0.35 and 0.96 log-change, depending on POTW) in incidence rates that is similar as to data presented before 1 May 2022 (between 0.65 and 0.96 log-change, depending on POTW), but a specific wastewater concentration corresponds to a lower observed incidence rate for data collected after 1 May 2022 than before 1 May 2022. This could be explained by fewer cases being reported through the public health case reporting system due to the increased use of at-home antigen tests, or decreased overall testing.

The change in the relationship between wastewater concentrations of SARS-CoV-2 RNA and reported laboratory-confirmed COVID-19 incidence rates is likely a result of decreased PCR test-seeking and availability, and increased availability of at-home COVID-19 antigen tests. It has been confirmed that PCR test seeking behavior has decreased as the availability of at-home antigen tests has increased^20^. Reduced test seeking behavior will bias the number of laboratory-confirmed cases downward but would not necessarily change the PCR testing positivity rate, so long as reduced test-seeking behavior is similar across all individuals including those who ultimately are infected and not infected with SARS-CoV-2. The similar relationship between SARS-CoV-2 RNA wastewater concentrations and laboratory-confirmed test positivity rate for data collected before and after 1 May 2022 would be consistent with this explanation.

It is important to note that there are other potential reasons that the relationship between SARS-CoV-2 RNA wastewater concentrations and laboratory-confirmed COVID-19 incidence rates could change over time. The duration and magnitude of SARS-CoV-2 RNA shedding via human excretions that enter wastewater might change as different SARS-CoV-2 variants circulate^32^. To date, there are limited quantitative, externally valid data (i.e., measurements reported as concentrations with units of copies per mass or volume of excretion) on SARS-CoV-2 RNA in excretions among individuals infected with different variants, particularly with the newer variants that have emerged over the last year ^33,34^. There is one study that inferred that the changing relationship between wastewater SARS-CoV-2 RNA concentration and case data between the Delta and Omicron surges in Arizona, USA was caused by changes in shedding^35^. Changes in severity of illness, or occurrence of asymptomatic infections may alter test seeking behavior, which would change case reporting, and yet is unrelated to the availability of at-home antigen tests.

It is highly likely that the relationship between wastewater concentrations and laboratory-confirmed COVID-19 incidence rates will continue to change as PCR test seeking and test availability changes. For example, an official end of the COVID-19 public health emergency in the United States may make both clinical laboratory and at-home testing unaffordable or more difficult to obtain for some individuals. The work herein suggests that, assuming SARS-CoV-2 RNA shedding remains relatively stable among those infected with the virus as different variants emerge, that wastewater concentrations of SARS-CoV-2 RNA can be used estimate COVID-19 cases as they would have been during the time when PCR testing availability and test seeking behavior were at a high (here, before 1 May 2022) using the historical relationship between SARS-CoV-2 RNA and case data.

## Data Availability

All data produced in this study are publicly available through the Stanford Digital Repository (https://doi.org/10.25740/xy132dg9314).

https://doi.org/10.25740/xy132dg9314

## Acknowledgements

We thank Allegra Koch and Amanda Bidwell for assistance with literature reviews, and Vikram Chan-Herur for comments on the final draft. We acknowledge California Department of Health for providing the sewershed-aggregated COVID-19 case data. Numerous people contributed to sample collection including Regional San Environmental Laboratory and Scientific Research Section Personnel (Sac), Payal Sarkar (SJ) and plant operations staff at San Jose-Santa Clara Regional Wastewater Facility, Lily Chan (OS), and the Oceanside plant operations and laboratory personnel. This study was performed on the ancestral and unceded lands of the Muwekma Ohlone people. We pay our respects to them and their Elders, past and present, and are grateful for the opportunity to live and work here.

