## Supporting Information for "Divergence of wastewater SARS-CoV-2 and reported laboratory-confirmed COVID-19 incident case data coincident with wide-spread availability of at-home COVID-19 antigen tests"

Supporting Information
for

### Divergence of wastewater SARS-CoV-2 and reported laboratory-confirmed COVID-19 incident case data coincident with wide-spread availability of at-home COVID-19 antigen tests

Alexandria B. Boehm[1]*, Marlene K. Wolfe[2], Bradley J. White[3], Bridgette Hughes[3], Dorothea Duong[3]

1. Department of Civil & Environmental Engineering, School of Engineering and Doerr School of Sustainability, Stanford University, 473 Via Ortega, Stanford, CA, USA 94305
2. Gangarosa Department of Environmental Health, Rollins School of Public Health, Emory University, 1518 Clifton Rd, Atlanta, GA, USA, 30322
3. Verily Life Sciences LLC, South San Francisco, CA, USA, 94080

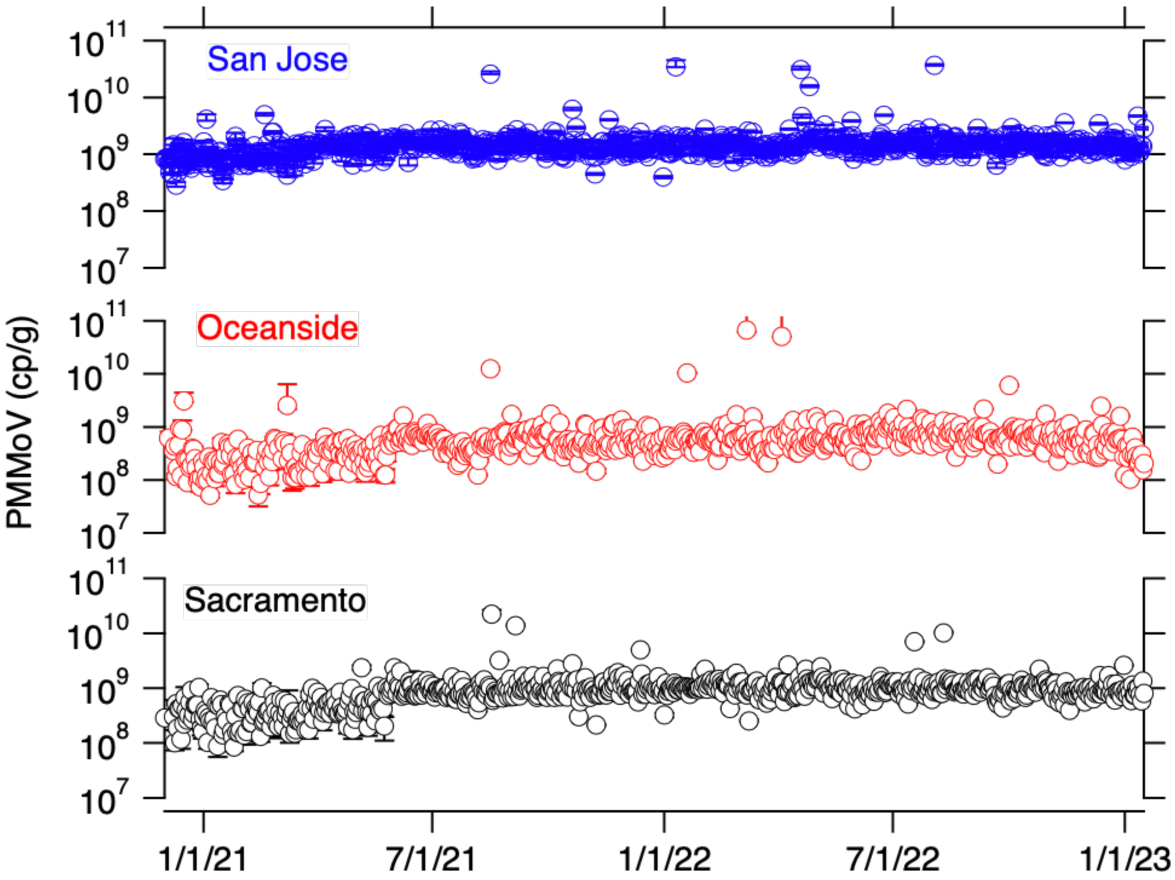


Figure S1. PMMoV concentrations in units of copies per gram dry weight and associated errors as represented by standard deviations. Errors are difficult to see in some cases because the size of the error bars are smaller than the symbol.

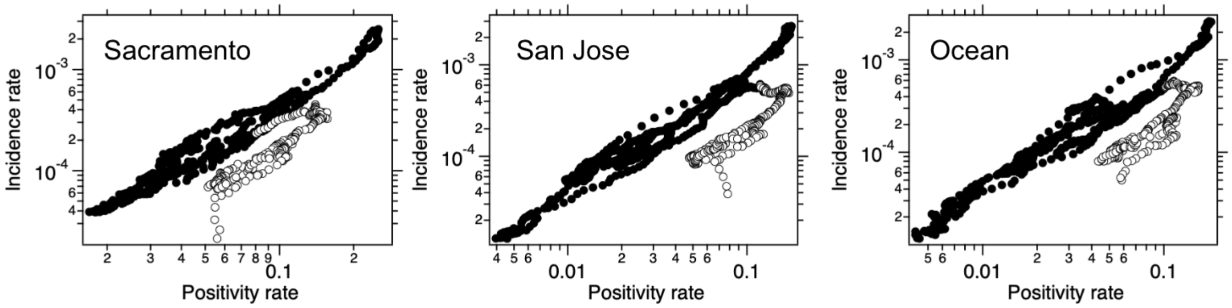


Figure S2. Relationship between incidence rate and positivity rate for the three POTWs. Here incidence rate and positivity rate are reported as fractions and the rates are the county-aggregated rates. Symbols colored black indicate data collected before 1 May 2022 and symbols colored white indicate data collected on or after 1 May 2022.

Table S1. Coefficients for the model Y = b + m*X + n*D + k*(D*X) presented as equation 1 in the text where X is $\log_{10}$-transformed positivity rate and Y is $\log_{10}$-transformed incidence rate. Adjusted R-square values and p-values for the model are provided, as well as the F statistic (and the degrees of freedom, DF). Coefficient values with standard errors are reported as well as p-values in parentheses. With Bonferroni corrections, p must be less than 0.0013 for alpha = 0.05. If n is significantly different from 0, then the intercept for the linear relationship between X and Y before and after 1 May 2022 is different and equal to b + n. If k is significantly different from 0, then the slope for the linear relationship between X and Y is different before and after 1 May 2022 and equal to m + k.

| POTW | b | m | n | k | Adjusted R2 | F-Statistic |
|---|---|---|---|---|---|---|
| SJ | -1.83±0.02 (0) | 1.27±0.01 (0) | -0.32±0.06 ($<10^{-7}$) | 0.19±0.05 (0.00016) | 0.92 ($<10^{-15}$) | 3188 on 3 and 773 DF |
| OS | -2.21±0.02 (0) | 1.09±0.01 (0) | -0.64±0.11 ($<10^{-8}$) | -0.17 ±0.09 (0.07) | 0.90 ($<10^{-15}$) | 2360 on 3 and 733 DF |
| SAC | -1.91±0.02 (0) | 1.43±0.02 (0) | -0.15±0.05 (0.0015) | 0.23±0.04 ($<10^{-6}$) | 0.93 ($<10^{-15}$) | 3548 on 3 and 765 DF |
